# The long-term success of mandatory vaccination laws at implementing the first vaccination campaign in 19^th^ century rural Finland

**DOI:** 10.1101/2020.12.14.20247577

**Authors:** Susanna Ukonaho, Virpi Lummaa, Michael Briga

## Abstract

In high income countries, childhood infections are on the rise, a phenomenon in part attributed to persistent hesitancy towards vaccines. To combat vaccine hesitancy, several countries recently made vaccinating children mandatory, but the effect of such vaccination laws on vaccination coverage remains debated and the long-term consequences are unknown. Here we quantified the consequences of vaccination laws on the vaccination coverage monitoring for a period of 63 years rural Finland’s first vaccination campaign against the highly lethal childhood infection smallpox. We found that annual vaccination campaigns were focussed on children up to 1 year old, but that their vaccination coverage was low and declined with time until the start of the vaccination law, which stopped the declining trend and was associated with an abrupt coverage increase of 20 % to cover >80 % of all children. Our results indicate that vaccination laws had a long-term beneficial effect at increasing the vaccination coverage and will help public health practitioners to make informed decisions on how to act against vaccine hesitancy and optimise the impact of vaccination programmes.

## Introduction

Infectious diseases are a major public health concern and socio-economic burden at all ages (1), but children are by far the most vulnerable to infections (2). In many low and middle-income countries, pertussis and measles remain among the top ten of all-cause mortality for children below age five (3,4). In high income countries, these infections are currently on the rise (5–11), despite the ready availability of vaccines.

One factor driving the increasing burden of childhood infections in high-income countries is vaccine hesitancy (5–11), i.e. partial or delayed acceptance or refusal of vaccination despite the availability of vaccines (12–14). Vaccine hesitancy is a key concern for public health institutions, because vaccination is our main tool to reduce and ultimately reach the local elimination of childhood infections (15). For the childhood infection measles and pertussis, reaching elimination requires a vaccination coverage of 95 and 98% of the population respectively (16,17), and such levels of vaccination coverage cannot be achieved with the current levels of hesitancy in high-income countries, especially for example, when there are spatial clusters of unvaccinated groups from which infections can re-emerge and spread towards other geographic regions (18).

One strategy to combat vaccine hesitancy is to adopt vaccination laws, i.e. make vaccination compulsory. Many states in the US and more recently in the European Union, e.g. in Italy, France and Germany in 2017, 2018 and 2020 respectively, have adopted vaccination laws which require children to be vaccinated against several childhood infections to be allowed to public schools or collective child services (19–21). These rules are enforced with fines (19,20), but children can be exempted for medical and in some states for various nonmedical reasons, which in the US resulted in a patchy vaccination coverage (22,23). Hence, to what extent vaccination laws are effective at increasing vaccination coverage is debated (24–29). So far, preliminary studies and short-term monitoring indicate that recent vaccination laws have been successful at generating a short-term increase in vaccination coverage in Italy and France (19,21) and in California, where a recent vaccination policy eliminated nonmedical exemptions from school entry requirements (30). However, to what extent this is a general result and whether the increase lasts on the long-term remain unknown.

In this study, we quantified the vaccination coverage and the long-term impact of vaccination laws on the vaccination coverage and its spatio-temporal dynamics during Finland’s first vaccination program against smallpox in the 19^th^ century. In brief, smallpox was one of the most lethal diseases in human history, affecting especially European and Native American populations (31–33). For example, in our study population in 18^th^ and 19^th^ century Finland, smallpox was a major cause of death in children, killing up to 15% of Finland’s population around 1800 before the introduction of vaccines (34–36).

Little is known on how the smallpox vaccination campaign was implemented in the 19^th^ century and to what extent it was successful (32), especially in rural areas where vaccine uptake remains low even today in many parts of the world (37). For the purpose of this study, we digitised a series of unique historical vaccination records (63 years, 1800 pages and over 46,000 individuals) maintained by the local church and the district doctor from the first vaccination campaign against smallpox in eight rural parishes of Finland (Fig. 1A, Fig. S1). This study has two aims. First, we described the seasonal and annual dynamics of the smallpox vaccination campaigns in rural Finland and which age categories were targeted. Knowing the age at vaccination is relevant for vaccine effectiveness: late vaccination diminishes protection against infection, thereby enabling the transmission of infection, while early vaccination can result in ineffective vaccination due to interference with maternal antibodies (38) in addition to considerable negative side effects (39,40). Second, we tested two hypotheses, namely: did the vaccination law cause (i) a long-term increase in *mean* vaccination coverage and (ii) a long-term decrease in *variance* in vaccination coverage. Increasing vaccination coverage and stabilizing its spatio-temporal variance are important because high variance can create pockets of susceptibles from which epidemics can re-occur (41). As vaccination behaviours tend to cluster within populations, highly vaccine hesitant communities may not reach herd immunity, thus posing a risk of disease outbreak (18). The high vaccine hesitancy in the 19^th^ century together with the rare long-term monitoring of vaccination coverage after the vaccination law provide a unique opportunity to quantify the long-term impact of vaccination laws in a high-opposition setting which is not yet possible in countries that recently implemented vaccination laws.

**Figure 1.**
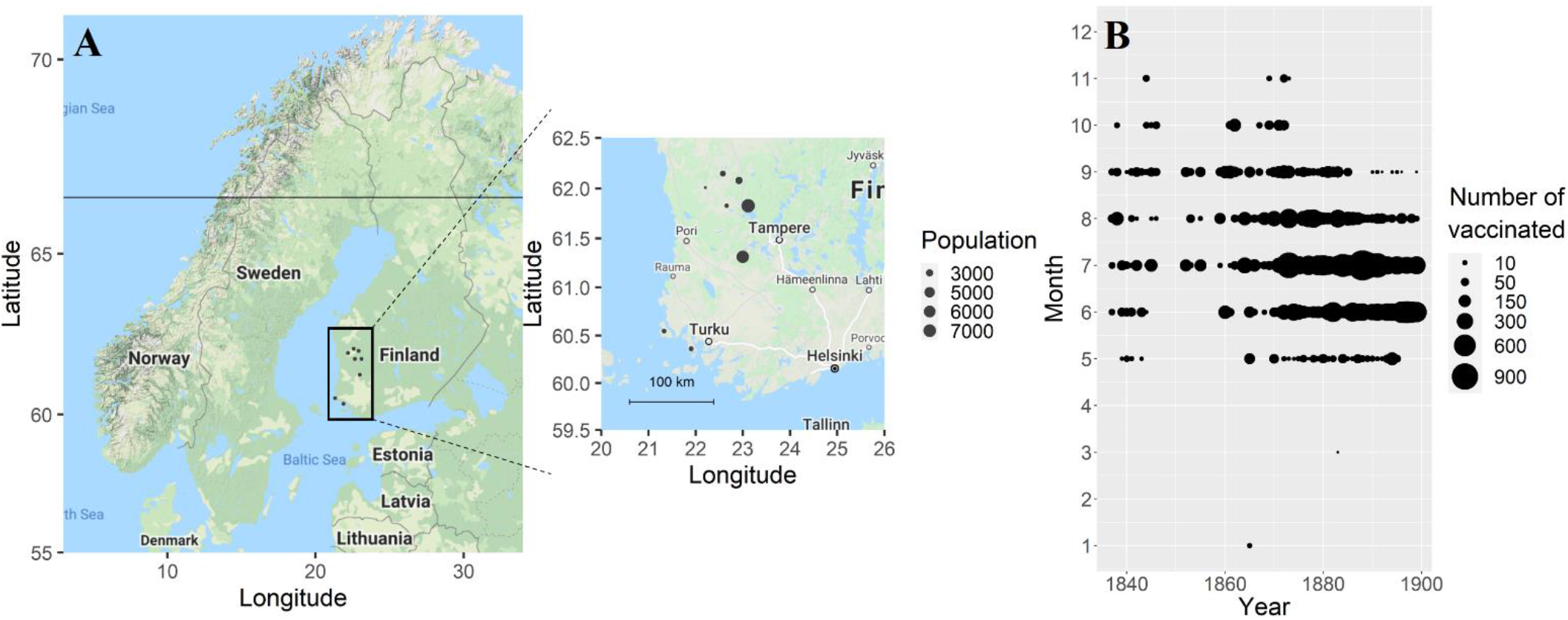
Geographical and temporal distributions of the data, showing (A) map of the eight rural parishes in south-west Finland with a map of Northern Europe on the left and a zoomed-in map of south-west Finland on the right, and (B) the predominant seasonal timing of annual vaccination campaigns occurring in summer.

## Methods

### Study population and data collection

Finland’s first vaccination campaign was met with high hesitancy from the public (42,43). It started in 1802, quickly after the development of world’s first vaccine by the British physician Edward Jenner in 1796 (44). Historical records describe that during our observation period, the distribution of vaccines improved gradually (42,43). In 1825 the country was divided into vaccination districts, which kept record of vaccinated and non-vaccinated citizens. Each district was supervised by a district doctor, who received annual reports from the vaccinators, who were physicians, clergymen and public officers. As the opposition against vaccines remained strong despite efforts to reassure the public, a mandatory vaccination law was adopted in 1883, which required parents to vaccinate children under the age of two, which was gradually enforced in 1885-1890 and onwards with a fine (42).

We photographed and digitised vaccination records manually from extensive church records held in Finnish national and provincial archives. These data include details of the villagers and their lives, that were by law required to be collected by local clergymen from 1749 onwards. The clergymen recorded the name, address, parents’ occupation, age, and the level of success of the vaccination (as indicated by reported reaction to the vaccine: effective, mild or no reaction) for each person receiving the vaccination at a given time, as well as previously vaccinated, absent or refused individuals (Fig. S1). Based on the accessibility and completeness of historical records, we obtained data from eight rural parishes (Honkajoki, Ikaalinen, Jämijärvi, Karvia, Kustavi, Parkano, Rymättylä, Tyrvää) from 1837 until 1899, but we lacked vaccination records from 1847-1851, 1854 and 1856-1858 for all parishes excluding Ikaalinen and Tyrvää and from Kustavi before 1862 and Rymättylä before 1865 (Fig. S2 & S3). These records include information on a total of 46,232 vaccinations, 26,328 and 19,935 records from the pre-mandatory and mandatory eras respectively (Table 1).

**Table 1.** Overview of the data, where we describe year, population and sample size, average number of vaccinated per year per parish, average timing and vaccination coverage per age category and interannual and interparish standard deviation (SD) and coefficient of variation (CV) of vaccination coverage for both pre-mandatory and mandatory eras. Children were grouped into the following age categories: under 1 year old (age <1) and between 1 and 5 years (from age 1 year until 5 years).

|  | Pre-mandatory era | Mandatory era |
| --- | --- | --- |
| Years | 1837-1882 | 1883-1899 |
| Average population size per parish | 2600 | 3700 |
| N vaccinated | 26328 | 19935 |
| Average n vaccinated per year per parish | 108 | 148 |
| Average [95CI] vaccination month | 7.40 ± 0.018<br>(mid-July) | 6.72 ± 0.011<br>(late June) |
| Average % of vaccinated under 1 year old | 68.1 | 87.5 |
| Average % vaccinated between 1 and 5 years | 16.3 | 10.3 |
| Interannual and -parish SD of vaccination coverage | 27.8 | 21.5 |
| Interannual and -parish CV of vaccination coverage | 40.9 | 24.6 |

In order to estimate the number of vaccinated children per age group, we gathered birth and death records for each parish from the national and provincial archives, the Genealogical Society of Finland and Finland’s Family History Association digitised archives. These records consisted of yearly births and deaths per age cohort (e.g. 0-1, 1-3, 3-5), which we used to calculate the number of individuals per age cohort per parish. We estimated the vaccination coverage based on the church records and total cohort sizes by calculating the yearly proportion of vaccinated per cohort and parish from 1837 to 1899. There were a few years with >100% vaccination coverage, which are likely the result of population movement. Birth records are unlikely the cause of vaccine coverage reaching over 100% as these have been validated by independent studies (45,46). Every five to ten years we had review tables of age-specific cohort sizes per parish from the historical records which confirmed our annually estimated cohort sizes.

## Statistical analyses

### Seasonality and age distribution of the vaccination campaign

The first aim of the study is to describe the occurrence and seasonal dynamics of the vaccination campaigns. To detect whether vaccination campaigns were seasonally recurring we conducted wavelet analysis. In brief, wavelet analysis decomposes time series data using functions (wavelets) simultaneously as a function of both time (year of observation) and period, with a period of 12 months indicating a yearly seasonally recurring pattern (47). We performed all statistical analyses with the software R (version: 3.6.1 (48)). For the wavelet analyses, we fitted a Morlet wavelet with the function ‘analyze.wavelet’ of the package ‘WaveletComp’ (49). To minimize the impact of any influential data points, we smoothened and detrended the data following standard protocol (49). We determined statistical significance by comparing the observed periodicity from the data with that of 1000 ‘white noise’ simulated datasets with [95%] significance level.

We then describe the age structure of the vaccination campaigns. For these analyses, we used a matrix with the number of vaccinated per age group per year and divided age into four groups (0-1, 1-3, 3-5, 5+). We tested for the preference towards children under one year old using non-parametric permutation tests with the function ‘independence_test’ of the package ‘coin’ (50) based on 10,000 permutations. We also describe the age at first vaccination for 872 individuals with exact birth dates.

### Spatio-temporal dynamics of vaccination coverage

To quantify the dynamics in vaccination coverage over time and across parishes and whether the vaccination law affected these dynamics, we used generalized linear mixed models (glms) and generalized additive mixed models (gamms). We ran four statistical models with as the dependent variable either: (a) the number of vaccinations, (b) the vaccination coverage, (c) the parish-level annual standard deviation (SD) in vaccination coverage and (d) the parish-level annual coefficient of variation (CV) in vaccination coverage (for explanation see below, Table S1A-D). In all these models the predictor variable was year standardised to the mean of zero and an SD of one.

We fitted glms with the functions ‘glm’, ‘lme’ and ‘gls’ respectively of the packages ‘stats’ (32) and ‘nlme’ (51). These gave consistent results and we here present the results using ‘glm’ for the number of vaccinated (Poisson distributed counts; model (a)), ‘lme’ for vaccination coverage (Gaussian distributed; model (b)) and ‘gls’ for parish-level SD and CV (Gaussian distributed with temporal autocorrelation, see below, models (c) and (d)). To quantify the statistical ‘significance’ of predictor variables, we compared the model fits on the data with the second order Akaike Information Criterion (AICc; (52,53)), using the function ‘AICc’ of the package ‘MuMIn’ (54). In brief, model selection ranks the models based on their AICc value, where better fitting models are indicated by lower AICc, models within 4 ΔAICc are considered plausible, and increasingly implausible up to 14 ΔAICc after which they are implausible (52,53).

To account for the fact that people are clustered into parishes, we included parish identity as a random intercept in model (b), the vaccination coverage analyses, which improved the model fit (ΔAICc=-29 in Table S1B). To have an idea of the importance of parish level differences, we reported the proportion of variance in the model explained by the random intercept and estimated the repeatability of parish-level differences with the function ‘rpt’ of the package ‘rptR’ (55). The repeatability is an intra-class correlation coefficient that captures the between-parish variance (by parish identity as a random intercept) relative to the total variance and ranges from zero (no parish-level differences) to one (all variance in the dependent variable is explained by parish-level differences). We estimated the 95% confidence intervals (95CI) around the repeatability based on 1000 bootstraps.

All model residuals were correctly distributed and fulfilled all other assumptions as checked with the function ‘simulateResiduals’ of the package ‘DHARMa’ (56). In model (b), there was however heteroscedasticity as identified with the functions (i) ‘testQuantiles’ from the package ‘DHARMa’ (56), and (ii) the Breusch-Pagan test performing a linear model using as dependent variable the squared value of the conditional residuals of the model (b) and standardised year as predictor variable (57). In model (b), we corrected for heteroscedasticity by including the variance power (‘varPower’) in the function ‘weights’ (58,59). We then tried to identify two sources of changes in heteroscedasticity by decomposing vaccination coverage into a parish-level component and an annual component. For the parish-level component, we used as the dependent variable the annual parish-level SD and CV in vaccination coverage (respectively models (c) and (d), Table S1 C & D). To identify whether there was an annual component in heteroscedasticity we used the Breusch-Pagan test as described above on the annual mean in vaccination coverage (excluding parish-level variance) and using pre-mandatory and mandatory era as a two-level categorical predictor.

We controlled for temporal autocorrelation following (60) by including standardised year as an auto-regressive factor of order 1 (corAR1). In models (a) number of vaccinated and (b) vaccination coverage, this worsened the model fit (ϕ=0.57±0.25, ΔAICc=+155 for model b in Table S1B), indicating no temporal autocorrelation, a conclusion also supported with the function ‘acf’ (ACF<0.1) of the package ‘stats’ (32). For these models, we hence presented the results without temporal autocorrelation, but analyses including temporal autocorrelation gave consistent conclusions (results not shown). Models (c) and (d), respectively parish-level SD and CV in vaccination coverage were autocorrelated and thus we included the corAR1 autocorrelation structure in these models.

To investigate whether there were abrupt changes in vaccination coverage or its spatio-temporal variance which coincided with the start of the vaccination law, we fitted threshold models following Douhard et al. (61) and Briga et al (62). In brief, for each model (a-d), we tested a series of threshold models, with one year interval between each threshold and identified the best fitting threshold model based on the model’s AICc value. To identify whether threshold models were appropriate at all, we compared the model fit of the best fitting threshold model with that of a linear model without threshold (Table S1 A-D). We identified the confidence interval around the threshold year by including all models within 4 ΔAICc of the best fitting threshold model (61,62), abbreviated as 4AICCI (Fig. S4 A, B, C & D). To confirm the results of threshold models, we performed all analyses also using generalized additive mixed models (gamms), with the function ‘gamm’ of the package ‘mgcv’, and to identify the year of change (inflection points) we used their derivatives with the function ‘fderiv’ of the package ‘gratia’ (63). All gamm analyses confirmed the conclusions obtained from the threshold models (Fig. S5 A-H).

## Results

### Seasonality and age distribution of the vaccination campaign

Here we present data covering eight rural parishes in South-West Finland (Fig. 1A), with 26,328 records (57%) during the 46 years of the pre-mandatory era from 1837 to 1882 (average 572 records per year) and 19,935 records (43%) during the 17 years of mandatory era from 1883 until 1899 (average 1173 records per year; Table 1).

During both eras, vaccination campaigns started in May or June, occurred predominantly in summer and often ended in August or September (Fig. 1B). This seasonal pattern was confirmed by wavelet analysis, where we observed a statistically significant annual vaccination periodicity (p<0.05), both on average over the whole observation period (p<0.05, Fig. S2A) and in the time dependent analyses (p<0.05, Fig. S2B).

We then investigated the age distribution of the people vaccinated during the campaigns. There was a clear preference towards children under age one, with in the pre-mandatory era on average 69% of the records being under age one, 21% between one and three years, 5.3% between three and five years and 4.5% five years old or older (χ^2^= 1143, p<0.0001, Fig. 2A). In the mandatory era, the preference towards one-year olds was 10% higher at 80% of the records and this increase was statistically significant (χ^2^= 39.73, p=2.9e-10, Fig. 2A). We did not observe a lower limit for the age at first vaccination (Fig. 2B): 872 of the 34,496 records (2.5%) of the children under the age of one were below the age of 3 months. Thus, there was a preference to vaccinate before age one which increased with time and with no apparent lower age limit.

**Figure 2.**
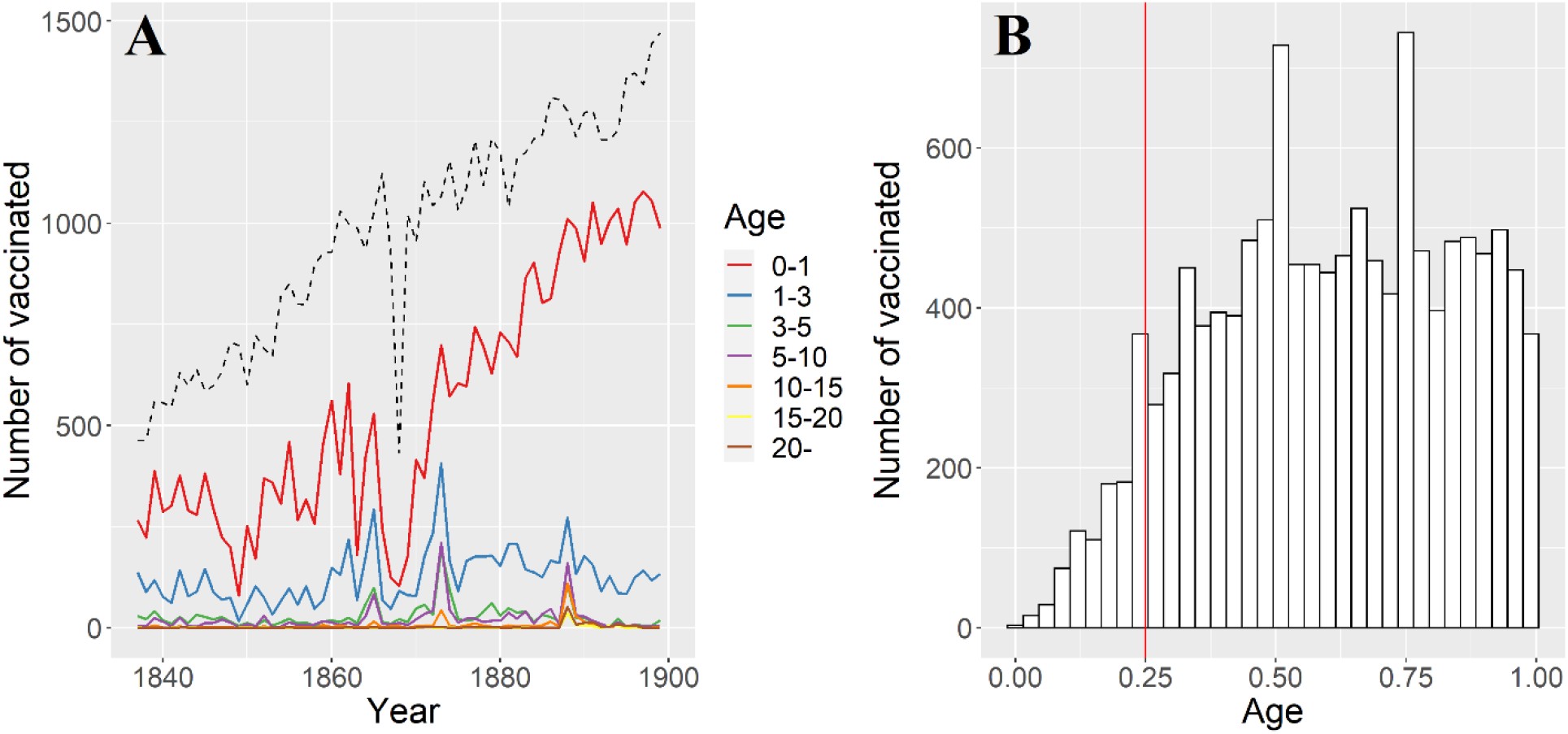
Smallpox vaccinations had a preference towards the youngest age groups (A) with no apparent minimum age (B). The number of vaccinated per age group (A) and age distribution for children under the age of one year (B). The dashed black line in (A) indicates the number of annual births in the eight study parishes. The red line in (B) indicates contemporary recommendation for age at first vaccination, which is 3 months in Finland for most vaccines. For smallpox, the recommended age at first vaccination is one year (39).

### Spatio-temporal dynamics of vaccination coverage

The number of vaccinations increased with time (Fig. 2A, Table S1A). Threshold models showed that this increase accelerated after the break point in 1871 (1871<4AICCI<1871; Fig. S4A) and this result was supported by gamm models (Fig. S5A&B). This break point coincided shortly after the drop in births during the famine years 1867-68. Because vaccination dynamics can be driven mostly by the dynamics of births (Fig. 2A), we studied birth-independent spatiotemporal dynamics of vaccinations by using vaccination coverage, i.e. the number of individuals vaccinated relative to the number of individuals in the age category and focused the analyses below on children until age one year.

We tested the hypothesis that the vaccination law increased the vaccination coverage. In the pre-mandatory era, the vaccination coverage was on average 68% (SD=27.8, 18.7<[95CI]<147) and in the mandatory era this increased with 20% to 88% (SD=21.5, 57.4<[95CI]<143; Fig. 3). Analysing these time series with threshold models showed an abrupt increase in vaccination coverage in 1882 (1882<4AICCI<1882; ΔAICc relative to a linear increase=-30.4; Table S1B; Fig. 4, Fig S4B). Before the threshold the vaccination coverage decreased with time, while after the threshold we observed an increase. Before the threshold, the 95CI of coefficient does not overlap with zero and hence is statistically significant, but after the threshold the 95CI overlaps with zero and hence is not significant (respectively: β_1_=−4.27, −8.35<[95CI]<-0.20, p=0.04; β_2_=9.22, −6.89<[95CI]<25.33; Table S1B). Gamm models supported the conclusions of threshold models with the gam derivative indicating the steepest increase in coverage in 1882 (Fig. S5C&D). Hence, the implementation of the vaccination law was associated with (i) a stop in declining coverage and (ii) an abrupt 20% increase in coverage which (iii) persistent on the long-term.

**Figure 3.**
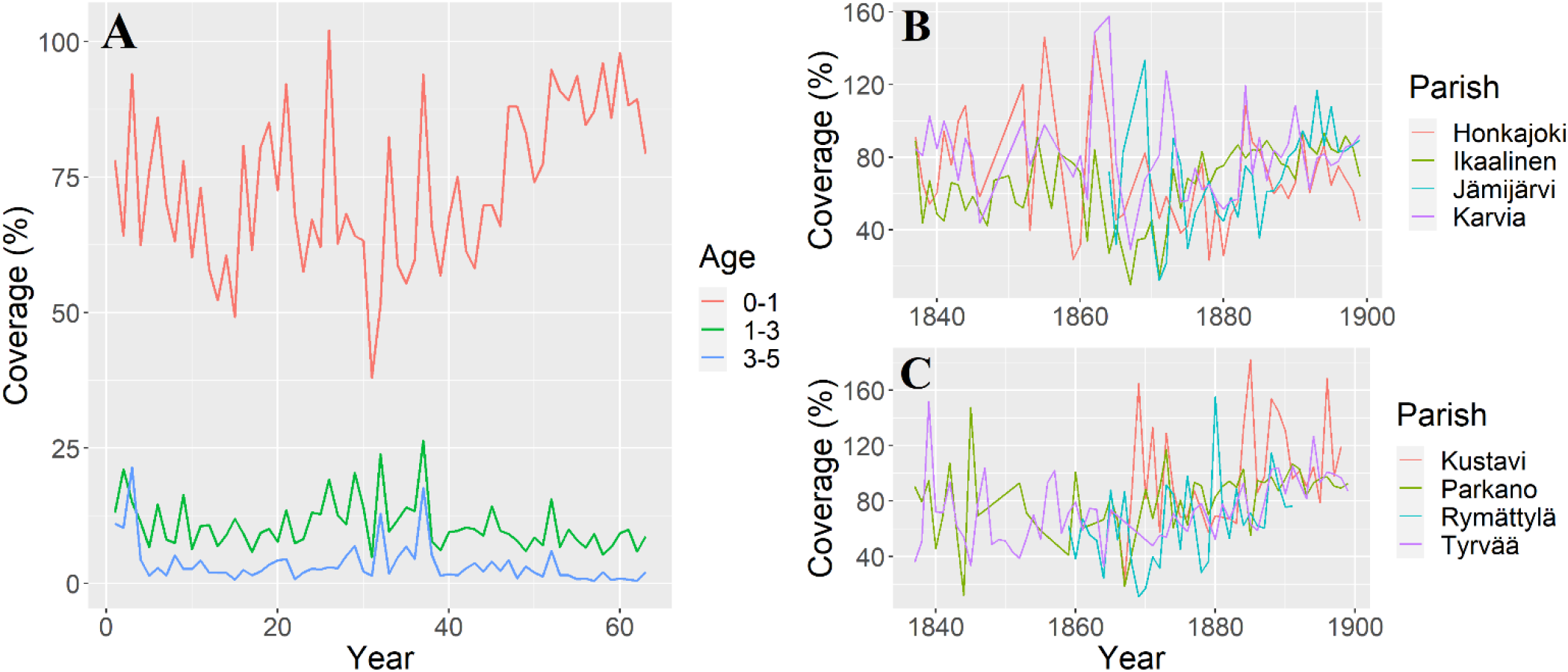
The vaccination coverage increased after the introduction of vaccination law in 1883 (A) together with the decrease in the variance between parishes, which, for illustrative purposes, are shown in two plots (B-C).

**Figure 4.**
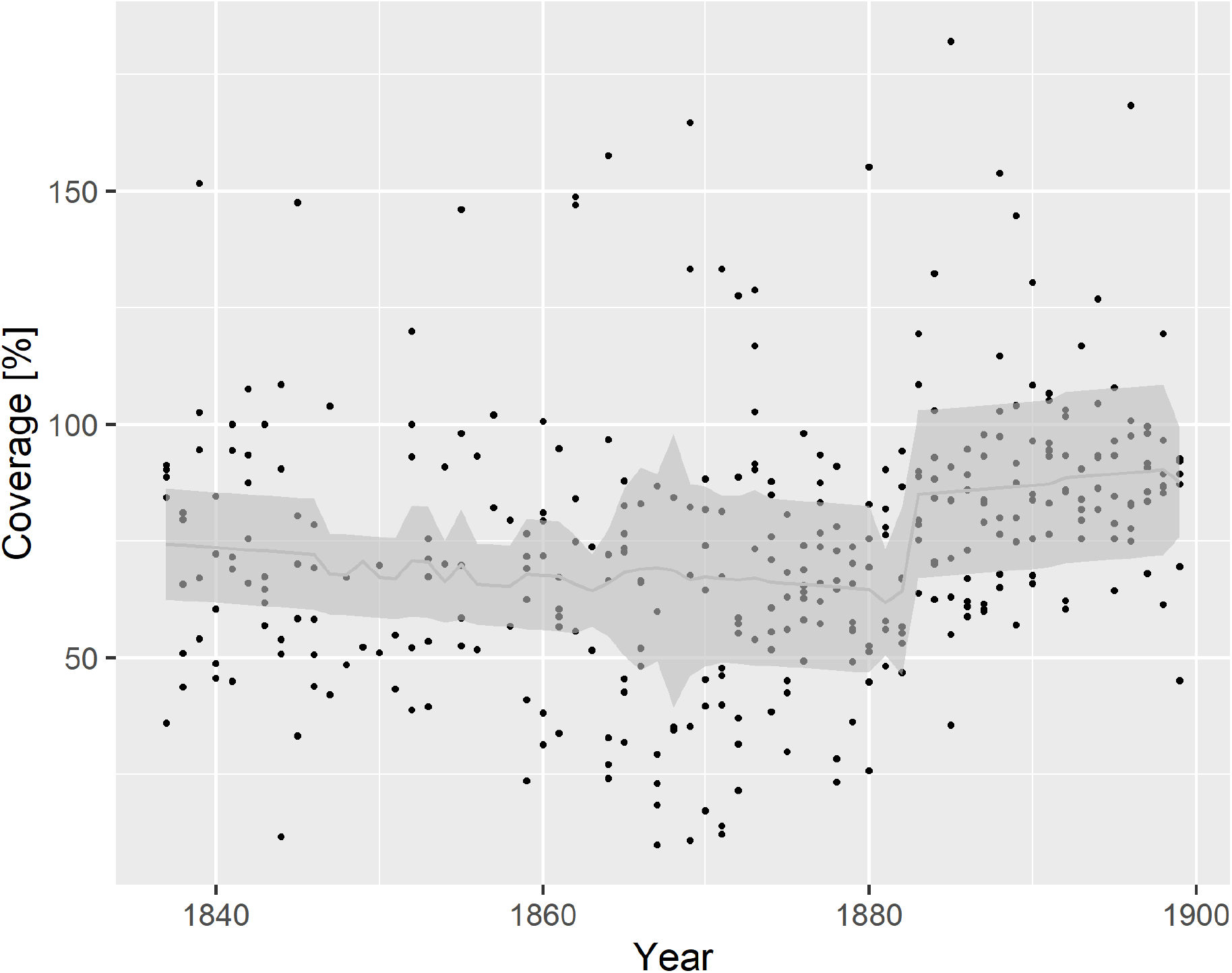
The temporal dynamics in vaccination coverage of children until age one year are best captured by a threshold model with an increase in vaccination coverage after1882. Before the threshold, the vaccination coverage decreases with time, while after the threshold there is an increase but the 95% CI of both coefficients overlap with zero and hence are not statistically significant. The predicted values are plotted as a light grey line against the black observed datapoints and the grey bands represent the [95%] confidence intervals.

We then tested whether the variance in vaccination coverage changed over time and Breusch-Pagan test indicated a decrease in variance (ΔAICc=-15.49). We tried to decompose whether this decrease in variance is due to a spatial (between parishes) and/or a temporal (between years) component. Vaccination coverage varied substantially and consistently between parishes: parish identity captured 9.0% of the variance in vaccination coverage during the pre-mandatory era which increased to 46% in the mandatory era (model in Table S1B) with a repeatability of respectively 0.083 (0.00<[95CI]<0.21) and 0.32 (0.069<[95CI]<0.55). The parish-level SD and CV in vaccination coverage decreased over time and showed an abrupt change around 1873 (SD: 1869<4AICCI<1876, CV: 1872<4AICCI<1874; Table S1C & S1D; Fig. 3B, Fig. S4C&D) and gamm models confirmed these conclusions (Fig. S5E-H). The interannual SD decreased as well with time, from 12.94 in the pre-mandatory era to 6.48 in the mandatory era (Breusch-Pagan test p=0.46). Hence the variance in vaccination coverage stabilized between parishes and over time, but the parish-level stabilization started approximately ten years before the introduction of the vaccination law.

## Discussion

Commonly preventable childhood infections are on the rise in many contemporary high-income countries following decreases in vaccination coverage, which has led to vaccination laws against infections such as measles and pertussis e.g. in Europe, many US states and Australia. However, we know almost nothing on the long-term effects of vaccination laws on vaccination coverage and its dynamics. In this study, we show that the Finland’s first vaccination campaigns in rural parishes were mainly executed during the summer, often in the middle of farming season and most of the vaccinated individuals were under the age of one year. We show an abrupt increase in vaccination coverage which coincided with the start of a vaccination law and persisted for at least 15 years. However, we also found differences in vaccination coverage between parishes, and although spatio-temporal variance in vaccination coverage decreased with time, this variance persisted and its sudden decrease ten years before the vaccination law appeared not to be associated with the vaccination law. Here we discuss three implications of our results for the impact of vaccination campaigns and vaccination policy.

First, low vaccination coverage in rural areas is a challenge that contemporary vaccination campaigns try to solve, especially in low and middle income countries (64–66). Increasing the vaccination coverage in these areas is important because spatial heterogeneity in vaccination coverage creates low vaccination clusters which can become a source of outbreaks that spread towards other areas (18). Our finding of repeatable parish-specific vaccination coverage show that vaccination coverage is a geographically local characteristic. The reasons behind the poor and fluctuating coverage in rural communities are complex (12,64,65), but at least two possibilities stand out. Firstly, remote parishes are often more difficult to access and possibly less well managed than more urban parishes (67). Secondly, people living in remote rural parishes might be less motivated or more hesitant to vaccinate (68,69). We observed a decline in the spatio-temporal variation in vaccination coverage, indicating improvements in the management of the vaccination campaign and reduction in inequity between the parishes. Although vaccination law probably attributed to this decline, it started a decade earlier and shortly after the famine years (18,67-68), which suggests that it is not directly associated with the vaccination law itself and is for example more likely due to improved accessibility in certain rural areas.

Second, one notable characteristic of the campaign was that children seemed to be vaccinated without lower age limit. Smallpox epidemics were still prevalent during the 19^th^ century and as vaccinators travelled to rural areas only once a year, early vaccination may have been the optimal solution at the time. However, we now know that early vaccination can be ineffective, for example through maternal blunting, where maternal antibodies of vaccinated mothers interfere with infant immune response and lower the infant’s responsiveness to vaccines (38), hence for example in contemporary societies a recommended age at first vaccination of at least three months for several childhood infections and one year for smallpox (39).

Finally, mandatory vaccinations have been one of the strategies considered to combat low vaccination coverage, but this approach has always been controversial and associated with opposition (70). Our results show an abrupt increase in the vaccination coverage at the start of the vaccination law. Similarly, short-term studies in contemporary societies observed immediate increases in vaccination coverage after the introduction of the vaccination laws (19,71) and our study suggested that the observed increases will persist on a longer term. Moreover, there were no major lethal smallpox outbreaks after 1882 (Fig. S6), reflecting that vaccination together with for example the general improvements in living conditions (72) diminished the lethal burden of smallpox.

Our study has several limitations. First, instead of vaccine hesitancy, vaccine delivery can be a major factor affecting vaccination coverage (73). In our study population, vaccine uptake likely improved at least in part due to better distribution and managing of the vaccination campaign. We cannot exclude sudden organisational changes of vaccination campaigns, but historical records indicate organisation improved gradually during our observation period (42,43). This is also indicated by the changes in CV and SD in vaccination coverage which occurred 10 years prior to the vaccination law, which probably reflects an improvement in Finland’s access to vaccines. Thus, in our study, the vaccination law and associated fines constitute just one factor that increased the vaccination coverage. Second, our study population was limited to eight rural towns from western-Finland. As vaccination campaigns may differ between cities or rural parishes, it is unclear to what extent we can extrapolate the characteristics of the vaccination campaign and the response to the vaccination law in time and space. Third, as parishes did not decline anybody wanting to get vaccinated, our estimates for the number of vaccinated also include temporary travellers from other parishes, which results in some data points with >100% vaccination coverage, indicating an overestimation of the vaccination coverage of the locals. Thus, vaccination campaigns were probably not quite as successful before or after the vaccination law as our estimates indicate.

The reasons behind changes in vaccine uptake remain complex and hence other measures than vaccination laws, such as training medical professionals about vaccines, actively recommending vaccines and good communication with the lay public are also needed to increase the vaccination coverage (29,74). Implementing vaccination laws with clear and consistent communication can help reach infection-specific thresholds for herd immunity and subsequent elimination (41,75). Our study indicates that vaccination laws will be an effective long-term tool in the public health battle to increase vaccination coverage and towards the elimination of childhood infections, even in socio-demographic contexts (rural areas, pre-health care periods) where vaccines are often met with higher reluctance than in urban areas.

## Data Availability

The data supporting the findings of our study is derived from three public domains: the National Archives of Finland, Finland’s Family History Association and the Genealogical Society of Finland. However, due to the nature of this study and personal information in these health records, supporting data are not publicly shared.

## Acknowledgements

Many thanks to Kimmo Pokkinen, Sara Itkonen and Tuija Koivisto for help with historical data, to the National Archives of Finland for allowing us to digitize and use the vaccination records and to Finland’s Family History Association and the Genealogical Society of Finland for access to the birth and death data. We are grateful to the Nordemics Consortium for useful discussions. For funding, we thank the valuable contributions from the Doctoral Programme in Biology, Geography and Geology, University of Turku, the Ella & Georg Ehrnrooth Foundation, the Academy of Finland (292368), ERC (CoG 648766) and NordForsk (104910). The authors declare no conflicts of interest. The data supporting the findings of our study are derived from three public domains: the National Archives of Finland, Finland’s Family History Association and the Genealogical Society of Finland.

## Supplementary material

**Figure S1.**
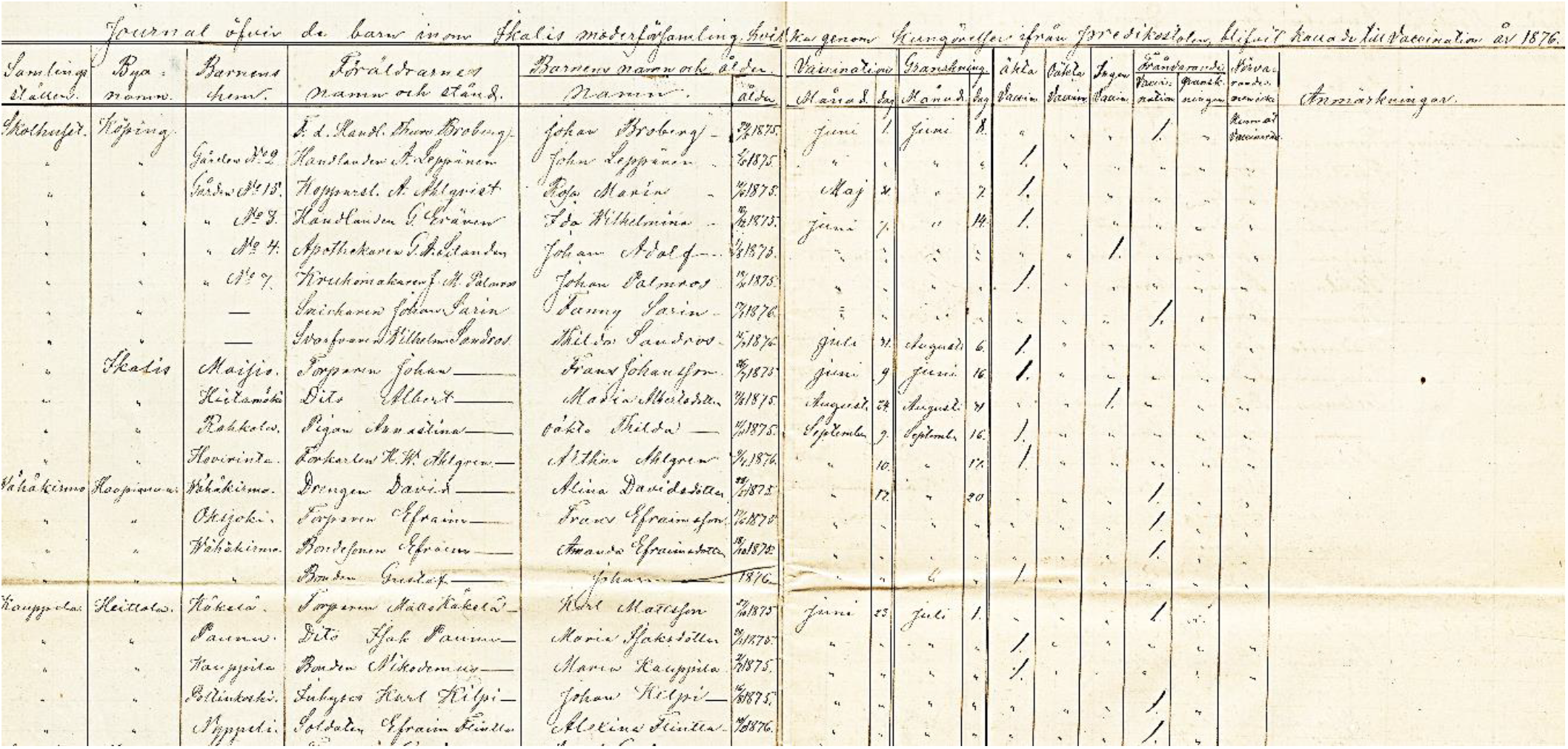
Illustration of Finnish historical vaccination records (Community: Ikaalinen, year: 1876, page 1). Columns represent (from left to right): I, ii, iii: Location, village and house; iv: Name and occupation of the parent (mother or father); v, vi: Names and birthdate of person called for vaccination; vii, viii: date of vaccination; ix, x: date of follow-up examination; xi-xiii: success of vaccination (successful, partial or no reaction); xiv-xv: absent from vaccination or follow-up examination. People who disagreed to vaccination are symbolised by ‘/’ or ‘x’ and do not have a date (not on this page); xvi: remarks and notes.

**Figure S2.**
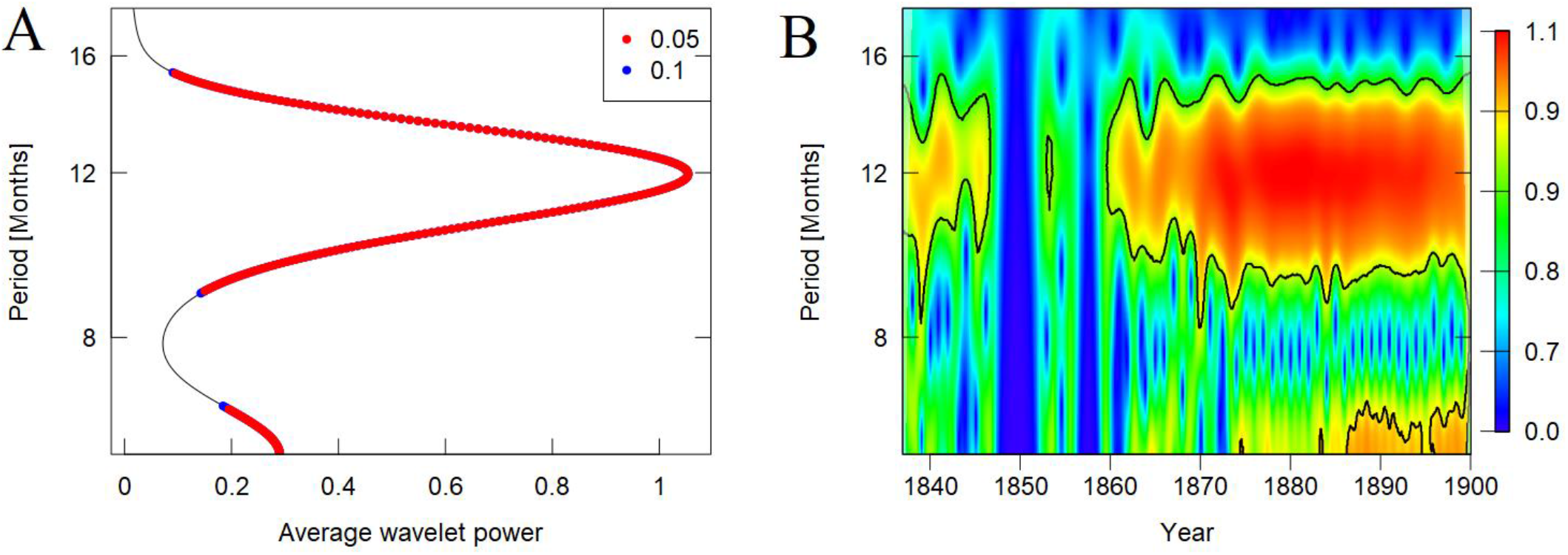
Wavelet periodogram (A) and wavelet power spectrum (B) for season at vaccination show a consistent average around one year from 1837 until 1899. A) A smaller peak can be observed at 6 months. Red dots represent significance at the 5% level and blue dots 10% level. The axis describe period in months on the y-axis and wavelet power levels on the x-axis. B) Legend describes wavelet power levels, where red indicates strong periodicity and green to blue signals weak periodicity. The blue gaps around 1850-1860 represent a gap in the data from all parishes at that time. The black line highlights the one-year mark in both plots.

**Figure S3.**
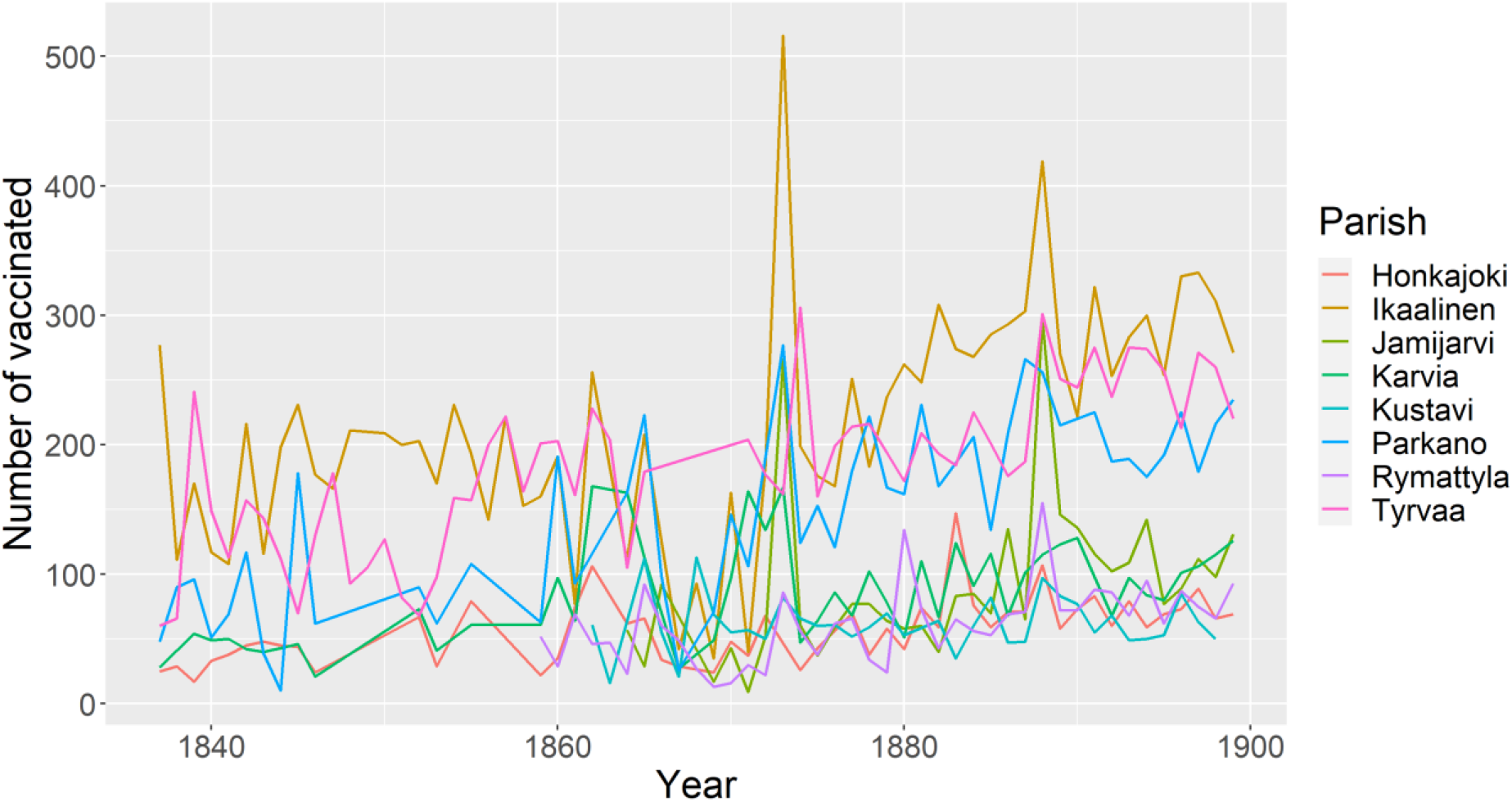
The number of vaccinated individuals increased towards the end of the 19^th^ century as monitored from 1837 to 1899.

**Table S1.**
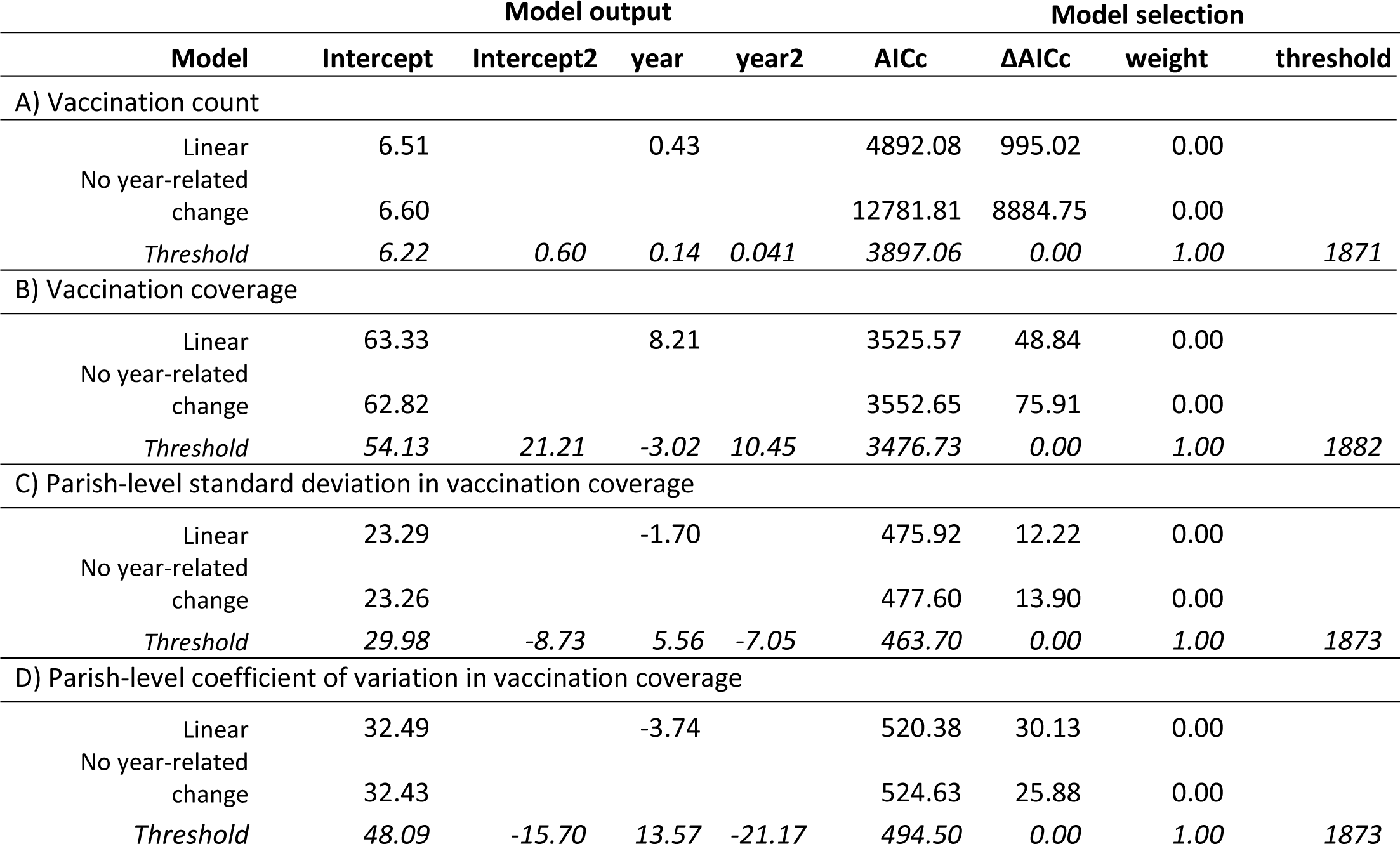
The model selection table indicated a quadratic increase in the number of vaccinated (count), vaccination coverage and coefficient of variation over time, but was not a significant predictor in standard deviation models. The table describes model type, model intercept, threshold intercept (Intercept2), scaled year coefficient, scaled threshold year coefficient (year2), period (before vs. after the law), model AICc, delta AICc, Akaike weight and threshold year for each dependent variable. The vaccination coverage model in B includes parish as a random variable. Best fitting models are indicated in italic.

**Figure S4.**
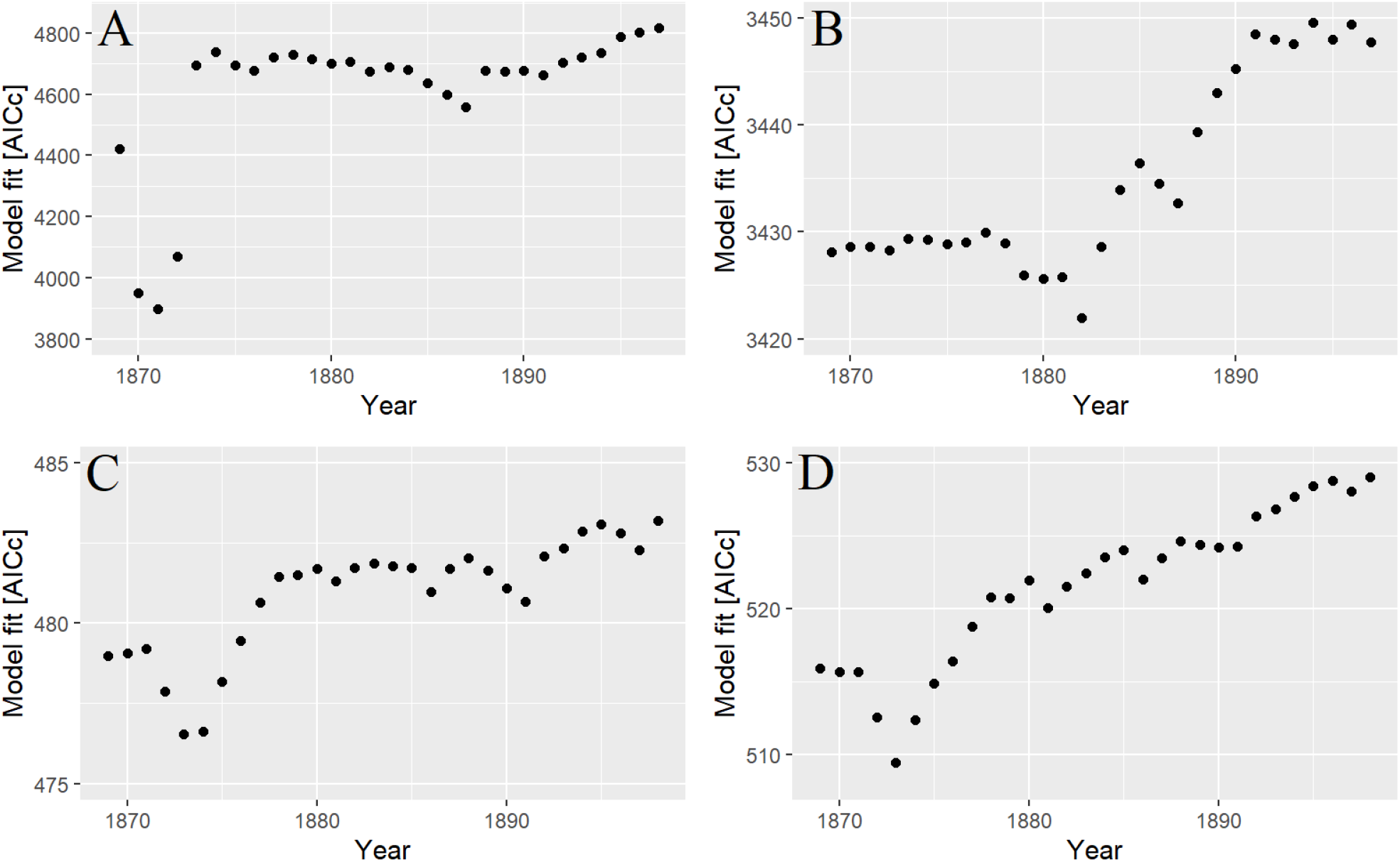
The number of vaccinated showed statistically significant threshold in 1871 (A, Table S1A), while the vaccination coverage showed a statistically significant threshold in 1882 (B, Table S1B). Both parish-level SD and CV models showed a statistically significant threshold in 1873 (C&D, Table S1C&D).

**Figure S5.**
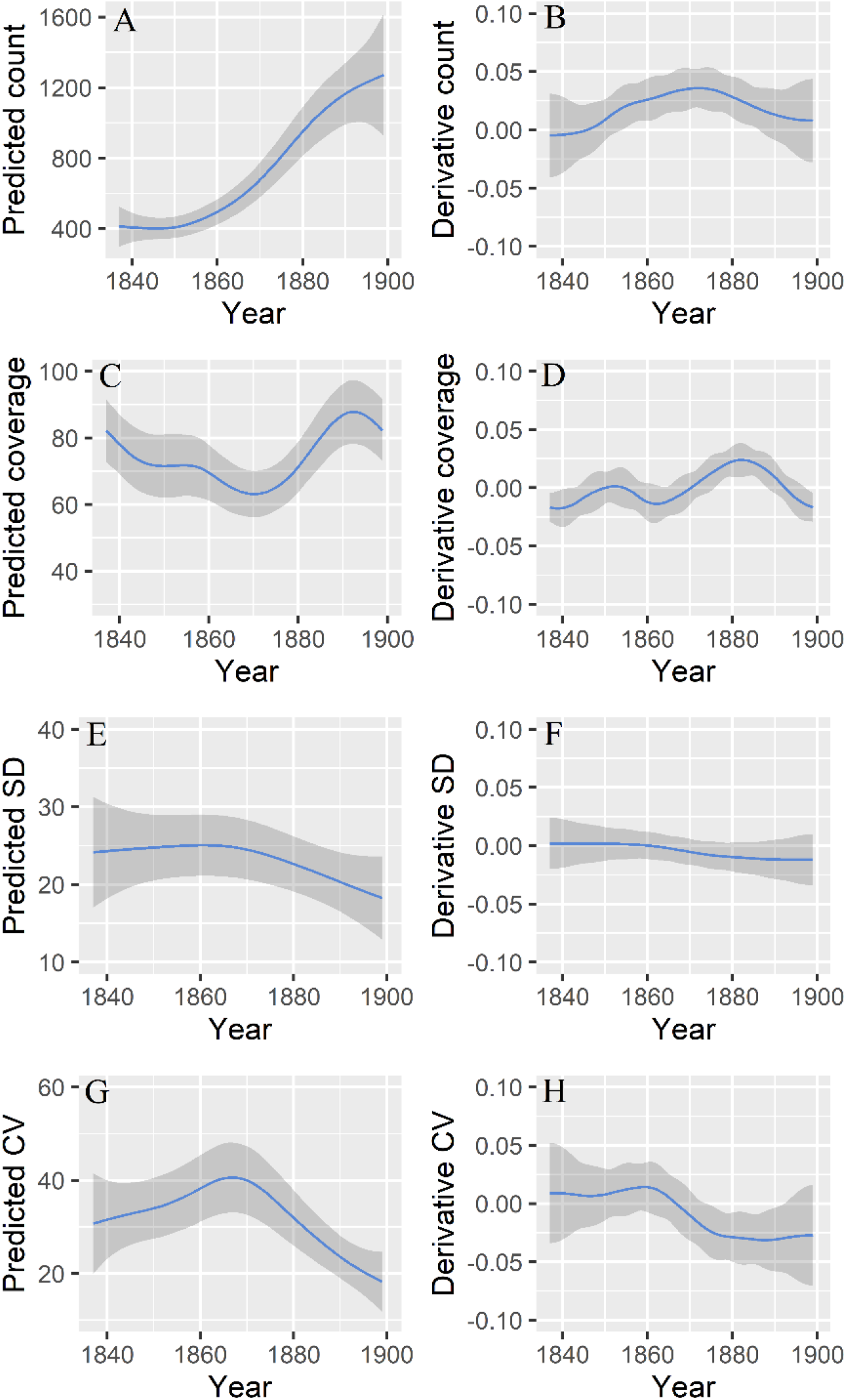
Gamm models show the steepest increase for number of vaccinated in 1871 (A) and vaccination coverage in 1882 (C), and for both parish-specific SD and CV a decrease starting from 1873 onwards (E&G). Shown here are the predicted values (A, C, E, G) and the derivatives of the GAMM models (B, D, F, H) for vaccination count (eq. model Table S1A; A&B), vaccination coverage (eq. model Table S1B; C&D) and parish-specific SD (eq. model table S1C; E&F) and parish-specific CV (eq. model Table S1D; G&H). Vaccination coverage models included parish identity as a random factor. All models used autocorrelation structure AR1 for year and estimated 95% confidence intervals (grey band) around the predicted slopes (blue line).

**Figure S6.**
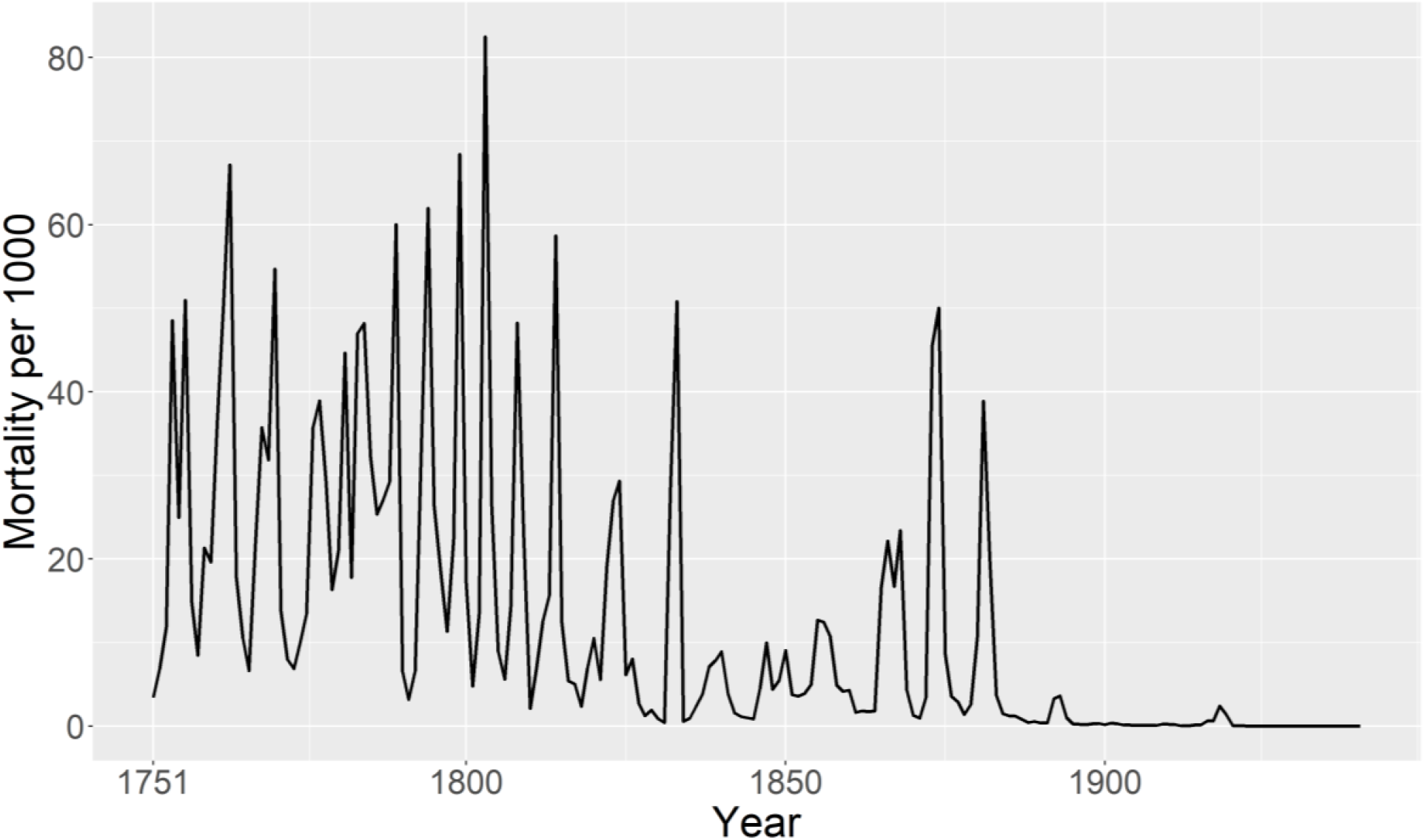
Smallpox mortality in Finland from 1751 to 1940. Original data from Pitkänen, Mielke & Jorde 1989 (Appendix Table 1).

## References

1. Morens DM, Folkers GK, Fauci AS. The challenge of emerging and re-emerging infectious diseases [published correction appears in Nature. 2010 Jan 7;463(7277):122]. Nature. 2004;430(6996):242–249. doi:10.1038/nature02759

2. Naghavi M, Abajobir A, Abbafati C, Abbas K, Abd-Allah F, Abera S-F, et al. Global, regional, and national age-sex specific mortality for 264 causes of death, 1980-2016: a systematic analysis for the Global Burden of Disease Study 2016 [published correction appears in Lancet. 2017 Oct 28;390(10106):e38]. Lancet. 2017;390(10100):1151–1210. doi:10.1016/S0140-6736(17)32152-9

3. Global Burden of Disease Child and Adolescent Health Collaboration, Kassebaum N, Kyu HH, et al. Child and Adolescent Health From 1990 to 2015: Findings From the Global Burden of Diseases, Injuries, and Risk Factors 2015 Study [published correction appears in JAMA Pediatr. 2017 Jun 1;171(6):602] [published correction appears in JAMA Pediatr. 2017 Oct 1;171(10):1019]. JAMA Pediatr. 2017;171(6):573–592. doi:10.1001/jamapediatrics.2017.0250

4. GBD 2019 Diseases and Injuries Collaborators. Global burden of 369 diseases and injuries in 204 countries and territories, 1990-2019: a systematic analysis for the Global Burden of Disease Study 2019 [published correction appears in Lancet. 2020 Nov 14;396(10262):1562]. Lancet. 2020;396(10258):1204–1222. doi:10.1016/S0140-6736(20)30925-9

5. Dabbagh A, Patel MK, Dumolard L, et al. Progress Toward Regional Measles Elimination - Worldwide, 2000-2016. MMWR Morb Mortal Wkly Rep. 2017;66(42):1148-1153. Published 2017 Oct 27. doi:10.15585/mmwr.mm6642a6

6. Omer SB, Salmon DA, Orenstein WA, deHart MP, Halsey N. Vaccine refusal, mandatory immunization, and the risks of vaccine-preventable diseases. N Engl J Med. 2009;360(19):1981–1988. doi:10.1056/NEJMsa0806477

7. Phadke VK, Bednarczyk RA, Salmon DA, Omer SB. Association Between Vaccine Refusal and Vaccine-Preventable Diseases in the United States: A Review of Measles and Pertussis [published correction appears in JAMA. 2016 May 17;315(19):2125] [published correction appears in JAMA. 2016 May 17;315 (19):2125]. JAMA. 2016;315(11):1149–1158. doi:10.1001/jama.2016.1353

8. Lo NC, Hotez PJ. Public Health and Economic Consequences of Vaccine Hesitancy for Measles in the United States. JAMA Pediatr. 2017;171(9):887–892. doi:10.1001/jamapediatrics.2017.1695

9. Olive JK, Hotez PJ, Damania A, Nolan MS. The state of the antivaccine movement in the United States: A focused examination of nonmedical exemptions in states and counties [published correction appears in PLoS Med. 2018 Jul 6;15(7):e1002616]. PLoS Med. 2018;15(6):e1002578. Published 2018 Jun 12. doi:10.1371/journal.pmed.1002578

10. Hotez PJ, Nuzhath T, Colwell B. Combating vaccine hesitancy and other 21st century social determinants in the global fight against measles. Curr Opin Virol. 2020;41:1–7. doi:10.1016/j.coviro.2020.01.001

11. Gardner L, Dong E, Khan K, Sarkar S. Persistence of US measles risk due to vaccine hesitancy and outbreaks abroad. Lancet Infect Dis. 2020;20(10):1114–1115. doi:10.1016/S1473-3099(20)30522-3

12. World Health Organisation. Report of the SAGE working group on vaccine hesitancy. Accessed October 1, 2014

13. Thomson A, Robinson K, Vallée-Tourangeau G. The 5As: A practical taxonomy for the determinants of vaccine uptake. Vaccine. 2016;34(8):1018–1024. doi:10.1016/j.vaccine.2015.11.065

14. Betsch C, Schmid P, Heinemeier D, Korn L, Holtmann C, Böhm R. Beyond confidence: Development of a measure assessing the 5C psychological antecedents of vaccination. PLoS One. 2018;13(12):e0208601. Published 2018 Dec 7. doi:10.1371/journal.pone.0208601

15. World Health Organisation. Global eradication of measles: report by the Secretariat. Accessed March 25, 2010.

16. Guerra FM, Bolotin S, Lim G, et al. The basic reproduction number (R0) of measles: a systematic review. Lancet Infect Dis. 2017;17(12):e420–e428. doi:10.1016/S1473-3099(17)30307-9

17. Kretzschmar M, Teunis PF, Pebody RG. Incidence and reproduction numbers of pertussis: estimates from serological and social contact data in five European countries. PLoS Med. 2010;7(6):e1000291. Published 2010 Jun 22. doi:10.1371/journal.pmed.1000291

18. Masters NB, Eisenberg MC, Delamater PL, Kay M, Boulton ML, Zelner J. Fine-scale spatial clustering of measles nonvaccination that increases outbreak potential is obscured by aggregated reporting data [published correction appears in Proc Natl Acad Sci U S A. 2021 Aug 10;118(32):]. Proc Natl Acad Sci U S A. 2020;117(45):28506–28514. doi:10.1073/pnas.2011529117

19. Gori D, Costantino C, Odone A, et al. The Impact of Mandatory Vaccination Law in Italy on MMR Coverage Rates in Two of the Largest Italian Regions (Emilia-Romagna and Sicily): An Effective Strategy to Contrast Vaccine Hesitancy. Vaccines (Basel). 2020;8(1):57. Published 2020 Jan 30. doi:10.3390/vaccines8010057

20. Neufeind J, Betsch C, Zylka-Menhorn V, Wichmann O. Determinants of physician attitudes towards the new selective measles vaccine mandate in Germany. BMC Public Health. 2021;21(1):566. Published 2021 Mar 22. doi:10.1186/s12889-021-10563-9

21. Lévy-Bruhl D, Fonteneau L, Vaux S, et al. Assessment of the impact of the extension of vaccination mandates on vaccine coverage after 1 year, France, 2019. Euro Surveill. 2019;24(26):1900301. doi:10.2807/1560-7917.ES.2019.24.26.1900301

22. Garnier R, Nedell ER, Omer SB, Bansal S. Getting Personal: How Childhood Vaccination Policies Shape the Landscape of Vaccine Exemptions. Open Forum Infect Dis. 2020;7(3):ofaa088. Published 2020 Mar 14. doi:10.1093/ofid/ofaa088

23. Wang E, Clymer J, Davis-Hayes C, Buttenheim A. Nonmedical exemptions from school immunization requirements: a systematic review. Am J Public Health. 2014;104(11):e62–e84. doi:10.2105/AJPH.2014.302190

24. Akmatov MK, Rübsamen N, Deyneko IV, Karch A, Mikolajczyk RT. Poor knowledge of vaccination recommendations and negative attitudes towards vaccinations are independently associated with poor vaccination uptake among adults - Findings of a population-based panel study in Lower Saxony, Germany. Vaccine. 2018;36(18):2417–2426. doi:10.1016/j.vaccine.2018.03.050

25. Maltezou HC, Theodoridou K, Ledda C, Rapisarda V, Theodoridou M. Vaccination of healthcare workers: is mandatory vaccination needed?. Expert Rev Vaccines. 2019;18(1):5–13. doi:10.1080/14760584.2019.1552141

26. Gualano MR, Olivero E, Voglino G, et al. Knowledge, attitudes and beliefs towards compulsory vaccination: a systematic review. Hum Vaccin Immunother. 2019;15(4):918–931. doi:10.1080/21645515.2018.1564437

27. Hadjipanayis A, Dornbusch HJ, Grossman Z, Theophilou L, Brierley J. Mandatory vaccination: a joint statement of the Ethics and Vaccination working groups of the European Academy of Paediatrics. Eur J Pediatr. 2020;179(4):683–687. doi:10.1007/s00431-019-03523-4

28. Vaz OM, Ellingson MK, Weiss P, et al. Mandatory Vaccination in Europe. Pediatrics. 2020;145(2):e20190620. doi:10.1542/peds.2019-0620

29. Musa S, Skrijelj V, Kulo A, et al. Identifying barriers and drivers to vaccination: A qualitative interview study with health workers in the Federation of Bosnia and Herzegovina. Vaccine. 2020;38(8):1906–1914. doi:10.1016/j.vaccine.2020.01.025

30. Nyathi S, Karpel HC, Sainani KL, et al. The 2016 California policy to eliminate nonmedical vaccine exemptions and changes in vaccine coverage: An empirical policy analysis. PLoS Med. 2019;16(12):e1002994. Published 2019 Dec 23. doi:10.1371/journal.pmed.1002994

31. Duncan SR, Scott S, Duncan CJ. The dynamics of smallpox epidemics in Britain, 1550-1800. Demography. 1993;30(3):405–423.

32. van Panhuis WG, Grefenstette J, Jung SY, et al. Contagious diseases in the United States from 1888 to the present. N Engl J Med. 2013;369(22):2152–2158. doi:10.1056/NEJMms1215400

33. Krylova O, Earn DJD. Patterns of smallpox mortality in London, England, over three centuries. PLoS Biol. 2020;18(12):e3000506. Published 2020 Dec 21. doi:10.1371/journal.pbio.3000506

34. Mielke JH, Jorde LB, Trapp PG, Anderton DL, Pitkänen K, Eriksson AW. Historical epidemiology of smallpox in Aland, Finland: 1751-1890. Demography. 1984;21(3):271–295.

35. Ketola T, Briga M, Honkola T and Lummaa V. Town population size and structuring into villages and households drive infectious disease risks in pre-healthcare FinlandProc. R. Soc. B. 2021; 288:20210356. 20210356. http://doi.org/10.1098/rspb.2021.0356

36. Ukonaho S, Lummaa V, Briga M. The long-term success of mandatory vaccination laws at implementing world’s first vaccination campaign in rural Finland [pre-print]. medRxiv. 2020. (doi: 10.1101/2020.12.14.20247577). Accessed December 16, 2020.

37. Ijarotimi IT, Fatiregun AA, Adebiyi OA, Ilesanmi OS, Ajumobi O. Urban-rural differences in immunisation status and associated demographic factors among children 12-59 months in a southwestern state, Nigeria. PLoS One. 2018;13(11):e0206086. Published 2018 Nov 5. doi:10.1371/journal.pone.0206086

38. Zimmermann P, Perrett KP, Messina NL, et al. The Effect of Maternal Immunisation During Pregnancy on Infant Vaccine Responses. EClinicalMedicine. 2019;13:21–30. Published 2019 Jul 26. doi:10.1016/j.eclinm.2019.06.010

39. Kempe CH. Studies smallpox and complications of smallpox vaccination. Pediatrics. 1960;26:176–189.

40. Cono J, Casey CG, Bell DM; Centers for Disease Control and Prevention. Smallpox vaccination and adverse reactions. Guidance for clinicians. MMWR Recomm Rep. 2003;52(RR-4):1-28.

41. Keeling MJ, Rohani P. Modeling infectious diseases in humans and animals. Princeton, 278 USA: Princeton University Press; 2011.

42. Björkstén J. Vaccinationens historia i Finland II. Helsingfors; 1908.

43. Björkstén J. Vaccinationens historia i Finland, I. Helsingfors; 1902.

44. Fenner F, Henderson D, Arita I, Jezek Z, Ladnyi I. Smallpox and its eradication. World Health Organization; 1988.

45. Pitkänen K. Deprivation and disease: Mortality during the Great Finnish Famine of the 1860’s. Helsinki: [Suomen väestötieteen yhdistys]: Tiedekirja [jakaja]; 1993.

46. Voutilainen M, Helske J, Högmander H. A Bayesian Reconstruction of a Historical Population in Finland, 1647–1850. Demography. 2020;57:1171–1192. https://doi.org/10.1007/s13524-020-00889-1

47. Cazelles B, Chavez M, Magny GC, Guégan JF, Hales S. Time-dependent spectral analysis of epidemiological time-series with wavelets. J R Soc Interface. 2007;4(15):625–636. doi:10.1098/rsif.2007.0212

48. R Core Team, R. A language and environment for statistical computing. R Foundation for Statistical Computing, Vienna, Austria; 2020. https://www.r-project.org/

49. Roesch A, Schmidbauer H. WaveletComp: Computational Wavelet Analysis. 2018.

50. Hothorn T, Bretz F, Westfall P. Simultaneous inference in general parametric models. Biom J. 2008;50(3):346–363. doi:10.1002/bimj.200810425

51. Pinheiro JC, Bates DM, DehRoy S, Sarkar D. nlme: Linear and Nonlinear Mixed Effects Models. R Core Team 2020. https://cran.r-project.org/package=nlme

52. Burnham KP, Anderson DR, Huyvaert KP. AIC model selection and multimodel inference in behavioral ecology: Some background, observations, and comparisons. Behavioral Ecology and Sociobiology 2011;65(1):23–35. https://doi.org/10.1007/s00265-010-1029-6

53. Burnham KP, Anderson DR. Model selection and multimodel inference: a practical information-theoretic approach. Springer-Verlag New York; 2002. https://doi.org/10.1007/b97636

54. Barton K. MuMIn: Multi-model inference. 2020. Available from: https://cran.r-project.org/package=MuMIn

55. Nakagawa S, Schielzeth H. Repeatability for Gaussian and non-Gaussian data: a practical guide for biologists. Biol Rev Camb Philos Soc. 2010;85(4):935–956. doi:10.1111/j.1469-185X.2010.00141.x

56. Hartig F. DHARMa: residual diagnostics for hierarchical (multi-level/mixed) regression models. 2020. https://cran.r-project.org/package=DHARMa

57. Breusch TS, Pagan AR. A Simple Test for Heteroscedasticity and Random Coefficient Variation. Econometrica 1979;47(5):1287–1294. https://doi.org/10.2307/1911963

58. Zuur AF, Ieno EN, Walker N, Saveliev AA, Smith GM. Mixed effects models and extensions in ecology with R. Springer-Verlag New York; 2009. https://doi.org/10.1007/978-0-387-87458-6

59. Pinheiro JC, Bates DM. Mixed-Effect Models in S and S-plus. Springer-Verlag New York; 2000. https://doi.org/10.1007/b98882

60. Woods S. Generalized Additive Models. An Introduction with R (2nd ed.). Chapman & Hall, Boca Raton, FI 2017. https://doi.org/10.1201/9781315370279

61. Douhard F, Gaillard J-M, Pellerin M, et al. The cost of growing large: costs of post-weaning growth on body mass senescence in a wild mammal. Oikos 2017;126(9):1329–1338. https://doi.org/10.1111/oik.04421

62. Briga M, Jimeno B, Verhulst S. Coupling lifespan and aging? The age at onset of body mass decline associates positively with sex-specific lifespan but negatively with environment-specific lifespan. Exp Gerontol. 2019;119:111–119. doi:10.1016/j.exger.2019.01.030

63. Simpson GL. gratia: Graceful ‘ggplot’-based graphics and other functions for GAMs fitted using “mgcv”. 2019. https://cran.r-project.org/package=gratia

64. Odusanya OO, Alufohai EF, Meurice FP, Ahonkhai VI. Determinants of vaccination coverage in rural Nigeria. BMC Public Health. 2008;8:381. Published 2008 Nov 5. doi:10.1186/1471-2458-8-381

65. Singh PK. Trends in child immunization across geographical regions in India: focus on urban-rural and gender differentials. PLoS One. 2013;8(9):e73102. Published 2013 Sep 4. doi:10.1371/journal.pone.0073102

66. Shikuku DN, Muganda M, Amunga SO, et al. Door - To - door immunization strategy for improving access and utilization of immunization Services in Hard-to-Reach Areas: A case of Migori County, Kenya. BMC Public Health 2019;19(1):1–11. https://doi.org/10.1186/s12889-019-7415-8

67. Gram L, Soremekun S, ten Asbroek A, et al. Socio-economic determinants and inequities in coverage and timeliness of early childhood immunisation in rural Ghana. Trop Med Int Health. 2014;19(7):802–811. doi:10.1111/tmi.12324

68. Wagner AL, Huang Z, Ren J, et al. Vaccine Hesitancy and Concerns About Vaccine Safety and Effectiveness in Shanghai, China. Am J Prev Med. 2021;60(1 Suppl 1):S77–S86. doi:10.1016/j.amepre.2020.09.003

69. Thomas TL, Caldera M, Maurer J. A short report: parents HPV vaccine knowledge in rural South Florida. Hum Vaccin Immunother. 2019;15(7-8):1666–1671. doi:10.1080/21645515.2019.1600986

70. Kennedy AM, Brown CJ, Gust DA. Vaccine Beliefs of Parents Who Oppose Compulsory Vaccination. Public Health Reports. 2005;120(3):252–258. doi:10.1177/003335490512000306

71. Partouche H, Gilberg S, Renard V, Saint-Lary O. Mandatory vaccination of infants in France: Is that the way forward?. Eur J Gen Pract. 2019;25(1):49–54. doi:10.1080/13814788.2018.1561849

72. van Wijhe M, McDonald SA, de Melker HE, Postma MJ, Wallinga J. Effect of vaccination programmes on mortality burden among children and young adults in the Netherlands during the 20th century: a historical analysis. Lancet Infect Dis. 2016;16(5):592–598. doi:10.1016/S1473-3099(16)00027-X

73. Smith PJ, Marcuse EK, Seward JF, Zhao Z, Orenstein WA. Children and Adolescents Unvaccinated Against Measles: Geographic Clustering, Parents’ Beliefs, and Missed Opportunities. Public Health Rep. 2015;130(5):485–504. doi:10.1177/003335491513000512

74. Larson HJ, Jarrett C, Eckersberger E, Smith DM, Paterson P. Understanding vaccine hesitancy around vaccines and vaccination from a global perspective: a systematic review of published literature, 2007-2012. Vaccine. 2014;32(19):2150–2159. doi:10.1016/j.vaccine.2014.01.081

75. Anderson RM, May R. Infcetious diseases of humans: dynamics and control. Oxford, UK: Oxford University Press; 1991.

